# Genome-wide Analysis of Copy Number Variation in Latin American Parkinson’s Disease Patients

**DOI:** 10.1101/2020.05.29.20100859

**Authors:** Elif Irem Sarihan, Eduardo Pérez-Palma, Lisa-Marie Niestroj, Douglas Loesch, Miguel Inca-Martinez, Andrea R. V. R. Horimoto, Mario Cornejo-Olivas, Luis Torres, Pilar Mazzetti, Carlos Cosentino, Elison Sarapura-Castro, Andrea Rivera-Valdivia, Elena Dieguez, Victor Raggio, Andres Lescano, Vitor Tumas, Vanderci Borges, Henrique B. Ferraz, Carlos R. Rieder, Artur Schumacher-Schuh, Bruno L. Santos-Lobato, Carlos Velez-Pardo, Marlene Jimenez-Del-Rio, Francisco Lopera, Sonia Moreno, Pedro Chana-Cuevas, William Fernandez, Gonzalo Arboleda, Humberto Arboleda, Carlos E. Arboleda-Bustos, Dora Yearout, Cyrus P. Zabetian, Timothy A. Thornton, Timothy D. O’Connor, Dennis Lal, Ignacio F. Mata, on behalf of the Latin American Research Consortium on the Genetics of Parkinson’s Disease (LARGE-PD)

**Author notes:** All the data for this manuscript was generated while IFM was affiliated at the VA Puget Sound and the University of Washington. Corresponding author: Ignacio F. Mata, PhD Lerner Research Institute R4-006, Cleveland Clinic Foundation, 9500 Euclid Ave. Cleveland, OH, 44195, USA.

## Abstract

**Background:** Parkinson’s disease is the second most common neurodegenerative disorder and affects people from all ethnic backgrounds, yet little is known about the genetics of Parkinson’s disease in non-European populations. In addition, the overall identification of copy number variants at a genome-wide level has been understudied in Parkinson’s disease patients.

**Objectives:** To understand the genome-wide burden of copy number variants in Latinos and its association with Parkinson’s disease.

**Methods:** We used genome-wide genotyping data from 747 Parkinson’s disease patients and 632 ancestry matched controls from the Latin American Research Consortium on the Genetics of Parkinson’s disease.

**Results:** Genome-wide copy number burden analysis showed no difference between patients vs. controls, whereas patients were significantly enriched for copy number variants overlapping known Parkinson’s disease genes compared to controls (Odds Ratio: 3.97 [1.69 - 10.5], *P* = 0.018). *PARK2* showed the strongest copy number burden, with 20 copy number variant carriers. These patients presented an earlier age of disease onset compared to patients with other copy number variants (median age at onset: 31 years vs. 57 years, *P* = 7.46 x 10^-7^).

**Conclusions:** We found that Parkinson’s disease patients are significantly enriched with copy number variants affecting known Parkinson’s disease genes. We also identified that out of 250 patients with early-onset disease, 5.6% carried a copy number variant on *PARK2* in our cohort. Our study is the first to analyze genome-wide copy number variants association in Latino Parkinson’s disease patients and provides insights about this complex disease in this understudied population.

## Introduction

Parkinson’s disease (PD) is the second most common neurodegenerative disorder, and the fastest growing cause of disability due to a neurological disorder in the world ^1,2^. PD is a multifactorial syndrome that is thought to be caused by the complex interaction of genetics, environmental factors, and aging ^3^.

Evidence for the genetic basis of PD has increased substantially over the past decades ^4–6^. The first causal gene for PD, *SNCA*, was discovered in 1997 ^7^, and its protein product (α-synuclein) was further shown to be a major component of Lewy bodies, the pathological hallmark of PD. Dominant pathogenic single nucleotide variants (SNVs) in *SNCA* ^8–11^, as well as copy number variants (CNVs), such as duplication or triplication of the entire gene with a clear dose effect, have been reported ^12–14^. The discovery of *SNCA* was followed by that of *PARK2* ^15^, where both pathogenic SNVs and CNVs are associated usually with autosomal recessive, early-onset form of the disease ^16^. Almost exclusively, genetic discoveries in PD have focused on SNVs, and studies on CNVs have been infrequent ^17–19^. CNVs in *PARK2, SNCA, PINK1, DJ1*, and *ATP13A2* (from more to less frequent) have been reported using a candidate gene approach ^20–22^, while no CNVs have been shown for *LRRK2*. To date, only two studies have investigated the role of CNVs in PD at a genome-wide level, including exclusively European and Ashkenazi Jewish individuals ^17,19^, with a sample size of 1672 and 432, respectively.

PD is a global disease affecting all ethnicities. Unfortunately, the majority of studies do not include individuals of non-European ancestry, creating a large gap in knowledge. This is especially true for Hispanics/Latinos (Box 1). Despite the fact that they are the largest and fastest growing ethnic minority in the US^23^, Hispanics/Latinos are critically underrepresented in most genetic studies ^24^. This is probably due to their complex admixed ancestry with influences primarily from European, Amerindian and African populations. In the US, the incidence and prevalence rates of PD among Hispanics are at least as high, if not higher than in non-Hispanic Whites, while the rates are lower for Asians and Blacks^25,26^. Yet, little is known about the genetics of PD in Hispanics/Latinos, especially the frequency and characteristics of CNVs. No genome-wide studies in this population have been performed to date.

#### Box 1

These terms are not interchangeable. The term “Latino’ refers to populations from Latin America (Mexico, most of Central and South America, and some islands in the Caribtiean), while the term “Hispanic” refers only to Spanish speaking populations, and does not include those Latin American countries where Portuguese or French is spoken. Since our cohort includes individuals from Brazil (who speak Portuguese), we refer to individuals from our cohort as Latinos, while most studies in the US only include His panics.

To address the lack of diversity in PD genetic studies and to understand the genetic architecture of PD in Latinos, we created the Latin American Research Consortium on the Genetics of PD (LARGE-PD) ^27^. For this study, we used genome-wide genotypes of 1,497 individuals from LARGE-PD. The aim of this study was to elucidate genomic structural changes, as well as assess the CNV burden in this cohort of Latino PD patients and controls.

## Methods

As part of our ongoing collaborative effort within LARGE-PD ^27^, we examined data from a total of 1,497 individuals (807 PD patients and 690 controls) recruited from nine different sites across the following five different countries: Peru (n = 721), Colombia (n = 351), Brazil (n = 227), Uruguay (n = 191), and Chile (n = 13). All patients were evaluated by a movement disorder specialist at each of the sites and met the UK PD Society Brain Bank clinical diagnostic criteria^28^. Controls were selected from ancestry matched individuals that did not have symptoms compatible with neurodegenerative disorders. All PD patients and controls provided signed informed consent according to the local ethical requirements of each site. All individuals were genotyped on Illumina’s Multi-Ethnic Global Array (MEGA) (Illumina, San Diego, CA, USA). A total of 1,779,819 markers were available before quality control (QC).

We performed an initial round of QC using PLINK 1.90 ^29^, based on single nucleotide polymorphism (SNP) genotype data for all samples and following established protocols described in Niestroj et al ^30^. Samples with a call rate < 0.96 or a discordant sex status were excluded. We filtered autosomal SNPs for low genotyping rate (> 0.98), case-control difference in minor-allele frequency (> 0.05), and deviation from Hardy-Weinberg equilibrium (HWE, P-value ≤ 0.001) before pruning SNPs for linkage disequilibrium (--indep-pairwise 200 100 0.2) using PLINK ^29^. 1000 Genomes population ^31^ was used as a reference for visual clustering of the Principal Component Analysis (PCA) to assess for population stratification.

For CNV calling, we focused only on autosomal CNVs due to the higher quality of CNV calls from non-sex chromosomes. A custom population B-allele frequency (BAF) file was generated as a reference before calling CNVs. Then, we created GC wave-adjusted Log R Ratio (LRR) intensity files for all samples and employed PennCNV ^32^ software to detect CNVs in our dataset.

We assessed cryptic relatedness using KING ^33^ software, and excluded individuals who were closely related (up to second degree) to another participant in our cohort by using the unrelated algorithm in KING. We performed an intensity-based QC to remove samples with low-quality data as previously described in Huang et al. ^34^ Following this step, all samples had a LRR standard deviation of < 0.27, absolute value of waviness factor < 0.03, and a BAF drift < 0.0014.

Called CNVs were removed from the dataset if they spanned < 20 markers, were < 20 Kb in length, and had a SNP density < 0.0001 (amount of markers/length of CNV). SNP density was not considered for CNVs spanning ≥ 20 markers and ≥ 1Mb in length, as larger CNVs are not likely to be artifacts. To ensure that only high-quality CNVs passed our filters, we implemented a quality score calculation for each CNV following the methods of Macé et al. ^35^, in which various CNV metrics are combined to estimate the probability of a called CNV to be a consensus call. Quality scores ranged from 0 (lowest) to 1 (highest) for duplications and similarly from 0 to −1 for deletions, and CNVs with quality scores between −0.5 to 0.5 were filtered out. A subset of final QC-passed CNVs were also inspected visually by five different investigators with expertise in the interpretation of BAF and LRR plots. CNVs were annotated for gene content using Ensembl ^36^ including gene name and the corresponding exonic coordinates in hg19 assembly using bedtools 2.27.0 ^37^.

We calculated CNV burden for PD using different categories to evaluate the relative contribution on PD risk: (1) the carrier status of overall CNV burden, including CNVs in non-genic regions (2) the carrier status of CNVs intersecting ‘any gene’ but none of the PD genes, (3) the carrier status of CNVs intersecting a list of “known PD genes”, and (4) the carrier status of large CNVs (≥ 1Mb in length). P values were adjusted with the false discovery rate (FDR) method to correct for multiple testing. For the overall CNV burden category, deletions and duplications were also analyzed separately. We selected 19 genes for the “known PD genes” category that were grouped as follows: six are well-established causal genes for PD (*LRRK2, PARK2, PARK7, PINK1, SNCA*, and *VPS35*), one is a susceptibility factor for PD with a large effect size (GBA), and 12 either result in a parkinsonian syndrome that sometimes overlaps PDptio or are putative causal genes for PD that have not be adequately validated (*ATP13A2, DNAJC6, DNAJC13, EIF4G1, FBXO7, GIGYF2, HTRA2, PLA2G6, RAB39B, SYNJ1, TMEM230*, and *VPS13C*) ^6,38,39^.

To assess for the difference in CNV burden between PD patients and controls, we fitted a logistic regression model using the “glm” function of the stats package ^40^ in R 3.6.0 ^41^. Cox-proportional hazards regression analyses and Kaplan-Meier curves were calculated using the survival package ^42^. For all burden analyses, odds ratios (OR), 95% confidence intervals (CIs), and significance were calculated. ORs were calculated by the exponential of the logistic regression coefficient. For Cox-proportional hazards regression, hazard ratios (HR) were calculated to allow for censored observations. Potential confounding variables were used as covariates and included age, sex, and the first five ancestry principal components for all regression models.

## Results

We had available data from a total of 1,497 individuals in LARGE-PD. We excluded 39 individuals due to relatedness, and 79 due to failing our intensity-based QC steps. Thus after QC, our final cohort included 1,379 individuals (747 PD patients and 632 controls) from Peru (n = 677), Colombia (n = 320), Brazil (n = 192), Uruguay (n = 177), and Chile (n = 13). There were more males in PD patients compared to controls (53.2% vs 33.1%, *P* < 0.001). Sample demographics are shown in Table 1. To visualize the ethnic composition of our cohort, we performed PCA using 1000 Genome populations ^28^ as a reference. Our samples overlapped strongly with the projection of Admixed Amerindian samples (AMR) (Supp. Fig. 1).

**Table 1:** Sample demographics. Number of PD patients and controls with their characteristics following QC steps.

|  | PD patients | Controls | *P* |
| --- | --- | --- | --- |
| Number of samples | 747 | 632 | ns |
| CNV carriers | 692 | 582 |  |
| Age (mean) | 62 | 56.6 | *** |
| Age onset (mean) | 54.4 | NA |  |
| Sex; Male (%) | 395 (53.2) | 209 (33.1) | *** |

PD: Parkinson's disease, CNV: copy number variant, \*\*\* : $P \leq 0.001$ , ns: non-significant, NA: not applicable.

The initial number of CNV calls was 249,101 including 176,462 deletions and 72,639 duplications. After all QC steps, including filtering by consensus quality score as described in the methods, the final number of high-quality CNVs was 8,412, including 5,155 CNVs in PD patients and 3,257 CNVs in controls. CNV analysis showed 1,274 of the samples (92.4%) carrying at least one QC-passed CNV. The length of the CNVs in the overall cohort ranged from 20 kilobases (Kb) to 3.4 megabases (Mb), with a median size of 52.4 Kb. CNV characteristics are shown in Supp. Table 1.

We applied logistic regression to compare the CNV burden in PD patients and controls on all categories defined earlier, adjusting P values for multiple testing (see methods for details). We found no significant difference in overall CNV burden (OR: 1.19 [0.78 - 1.8], *P* = 0.64), CNVs in any gene (OR: 1.07 [0.81 - 1.4], *P* = 0.77), and large CNVs that are ≥ 1Mb in length (OR: 1.46 [0.82 - 2.65], *P* = 0.4) (Fig. 1). Interestingly, nine PD patients and six controls carried a duplication that was ≥ 1Mb on chromosome 11 (*P* = 0.8), which did not overlap with any known gene region. We also calculated overall CNV burden independently in duplications (OR: 1.36 [1.05 - 1.77], *P* = 0.06) and deletions (OR: 0.98 [0.74 - 1.29], *P* = 0.89), and neither were statistically significant.

**Figure 1:**
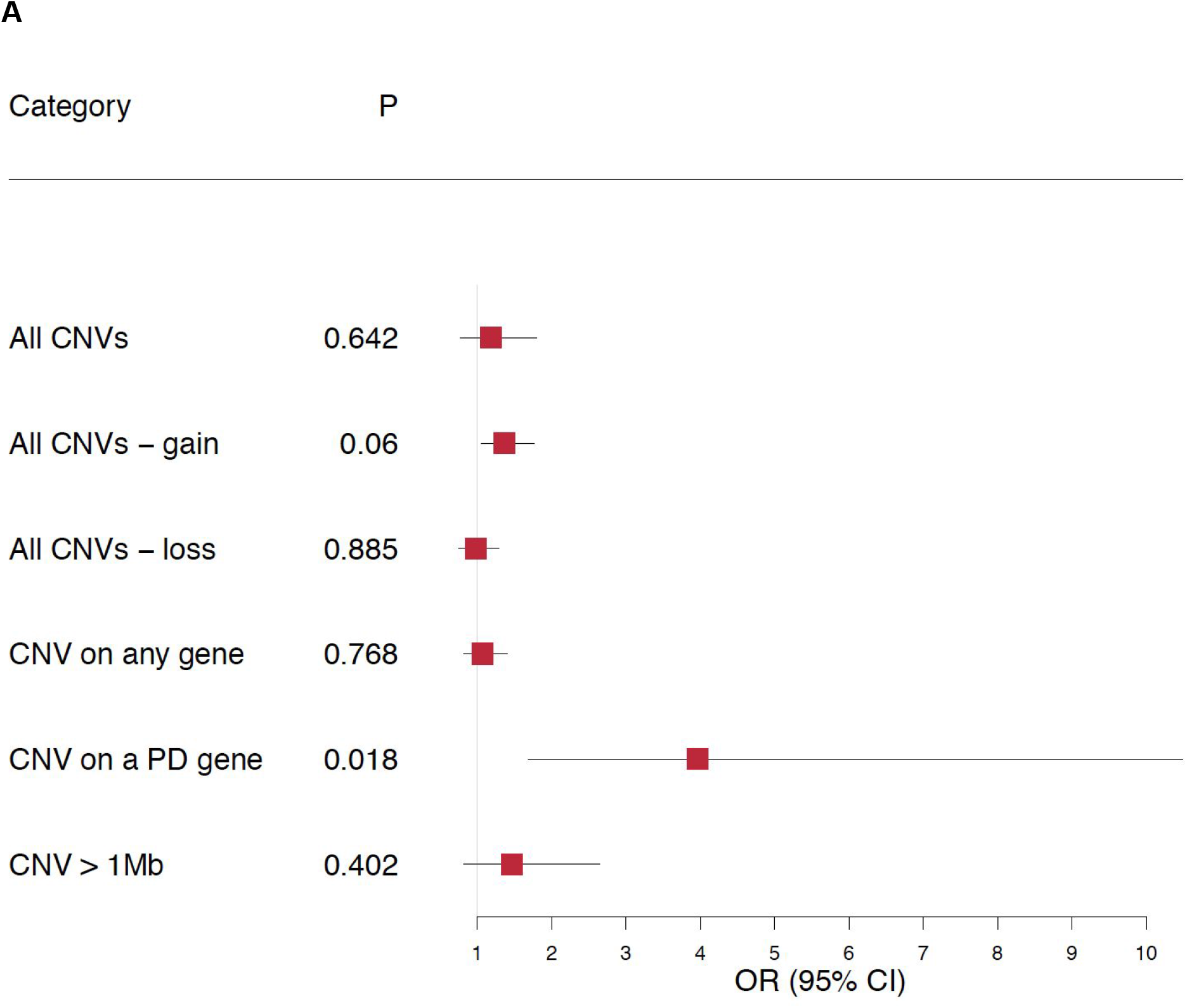
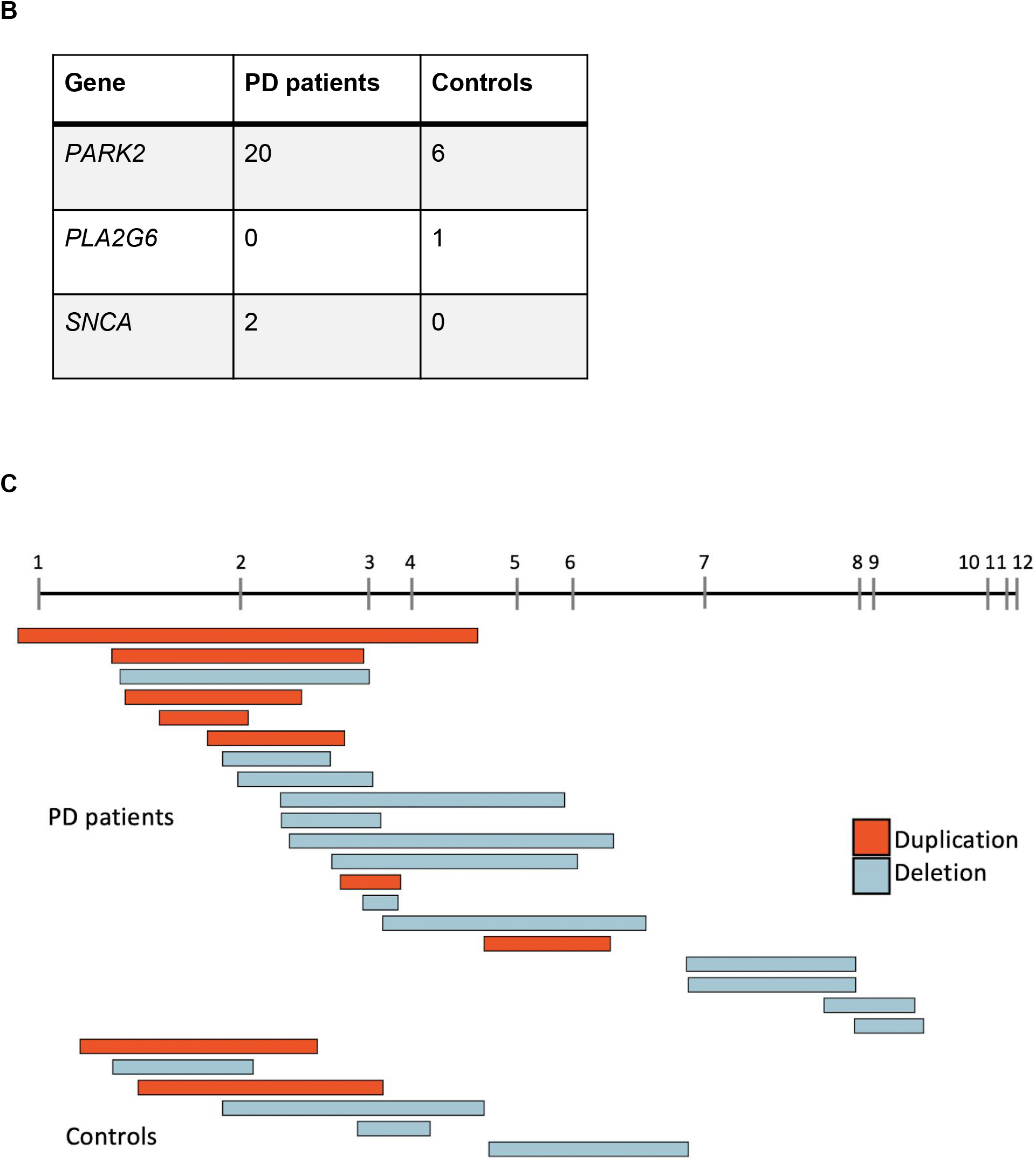
(A) Forest plot showing the CNV burden compared between PD patients and controls. Odds ratios (ORs) and P values were calculated using a logistic regression for CNVs corrected with age, sex, and first 5 components of PCA. P values were adjusted with FDR for multiple testing. ORs > 1 indicates an increased risk for PD per unit of CNV burden. (B) Table showing number of CNV carriers in any of the 19 known PD genes. (C) Visualization of CNVs on *PARK2*.

We then explored CNVs on genomic regions that were previously associated with typical PD and other parkinsonian phenotypes, and we found that PD patients were significantly enriched with CNVs overlapping these genes (OR: 3.97 [1.69 - 10.5], *P* = 0.018) (Fig. 1). This finding was largely driven by CNVs on *PARK2* in 20 patients, followed by two patients with a CNV on *SNCA*, compared to six controls carrying a CNV on *PARK2* and none on *SNCA*. In addition one control had a CNV on *PLA2G6* (Fig. 1).

To assess whether PD patients carrying a *PARK2* CNV in our cohort had an earlier age at onset (AAO), we performed a Mann-Whitney test and compared patients that carry a CNV overlapping *PARK2* to those carrying a CNV overlapping any other gene. The median AAO for patients with a CNV overlapping *PARK2* was 31 years old, while that for patients with a CNV overlapping any other gene was 57 years old (*P* = 7.46 x 10^-7^). To further investigate the CNV burden in early-onset PD (EOPD) patients, we performed a subset analysis in which cases with an AAO < 50 years old were compared to controls. Again, we observed a significant enrichment in CNVs on known PD genes in patients with EOPD (OR: 4.91 [1.92 - 13.68], *P* = 0.006). This result was also driven by CNVs on *PARK2*.

Kaplan-Meier estimates of AAO showed that individuals carrying a CNV on a known PD gene had significantly earlier onset of symptoms compared to individuals with other or no CNVs (log-rank test, *P* < 0.001) (Fig. 2). Using a Cox proportional-hazards regression analysis with age, sex, and the first five ancestry principal components included as covariates, we found that the effect of carrying a CNV on a known PD gene on the hazard of AAO was highly significant (HR: 2.42 [1.57 - 3.71], *P* = 5.70 x 10^-5^). We also assessed AAO in PD patients only, comparing CNV carriers on a known PD gene to PD patients with other or no CNVs, and found that having at least one CNV results in earlier onset of symptoms. (HR: 1.92 [1.22 - 3.02], *P* < 0.001) (Supp. Fig. 2).

**Figure 2:**
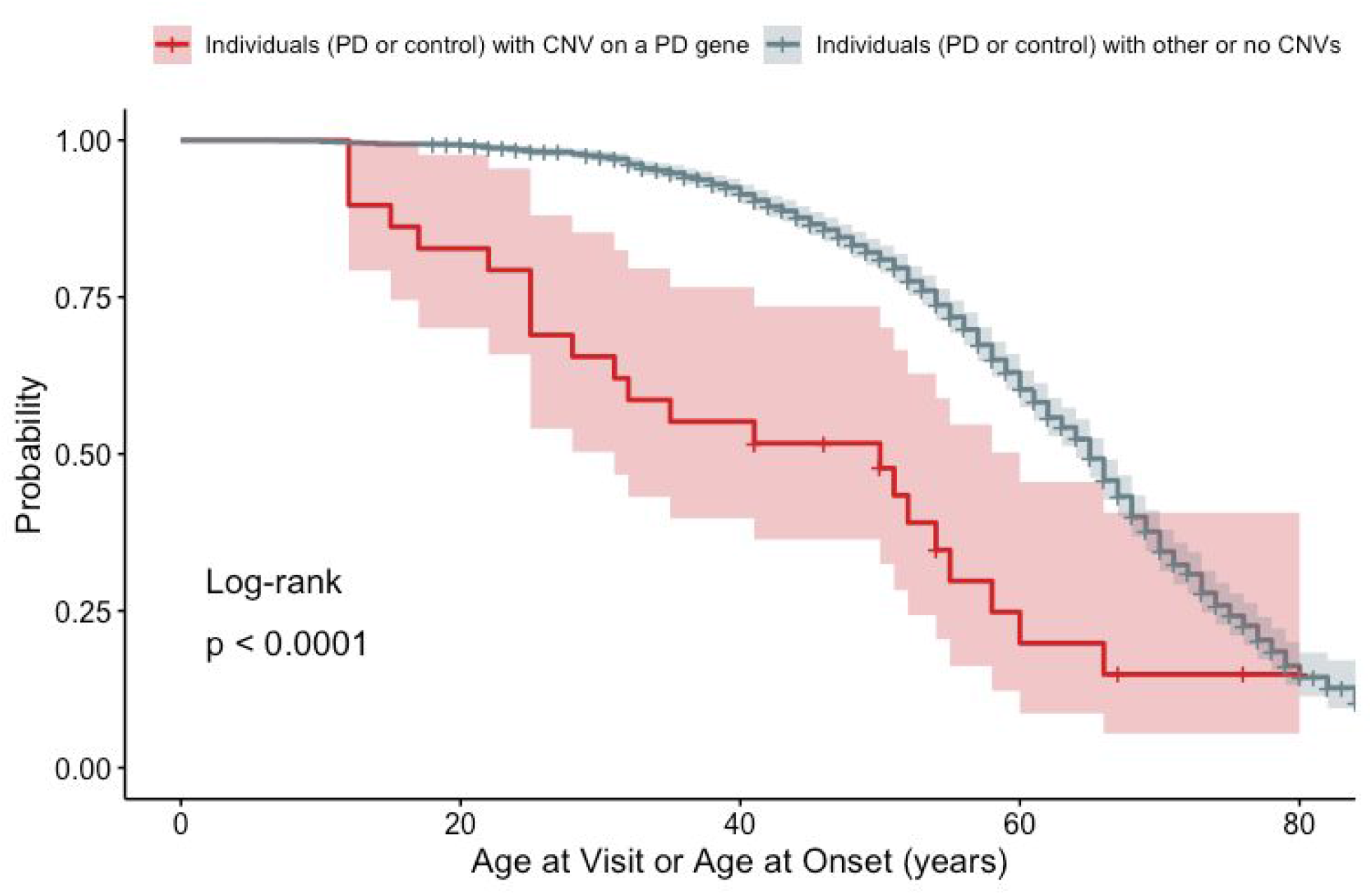
Kaplan-Meier estimates of individuals (PD patients and controls) carrying a CNV on a PD gene and individuals with other or no CNVs. Controls are censored observations since it is only known that they did not develop PD up to the age of their last visit. Probability: probability of not having symptoms associated with PD. Age at Visit or Age at Onset: time to onset of PD symptoms for cases and time to last visit for controls. Highlight around the curves shows 95% confidence intervals.

## Discussion

Here, we present a genome-wide characterization of CNVs in a cohort of Latino PD patients and controls from LARGE-PD ^27^. We analyzed genotypes of 1,497 individuals on the same platform and analyzed all samples with the same CNV calling and quality control pipeline. We used ancestry matched controls for the interpretation of CNVs detected in PD patients. This is particularly important considering that the data for Latino population frequency of CNVs is limited, especially in neurologically healthy adults ^43,44^. We assessed the CNV burden for different categories and observed an increased burden of CNVs overlapping known PD genes in PD patients. We identified 22 patients that carried CNVs overlapping two established PD genes (*PARK2* & *SNCA*), and found that 14 of these patients had a disease AAO < 50 years. The median AAO for patients with a CNV overlapping *PARK2* was almost 20 years earlier than that of other patients, in agreement with the literature ^45–47^.

*PARK2* mutations are the most common genetic cause for EOPD ^5,15,48^, but an important caveat is that this information is mostly derived from studies in populations of European ancestry. The frequency of all *PARK2* pathogenic variants (CNVs and SNVs) in EOPD patients with European ancestry ranges from 49% in familial cases, to 15-18% in isolated patients, while the frequency of carrying a CNV in *PARK2* in isolated EOPD patients is approximately 10%^45,49,50^. Some studies suggest that alterations in *PARK2* are more frequent in Hispanic populations. One study showed a 2.7 fold increase for carrying any *PARK2* alteration and a 2.8 fold increase for carrying a heterozygous mutation in Hispanic EOPD patients (N = 77) compared to White non-Hispanics, and the frequency of *PARK2* CNVs in Hispanic PD patients in this cohort was 6.4% ^48^. In another study examining Mexican-mestizo EOPD patients (N = 63), the frequency of *PARK2* CNVs was found to be 50%, and 18% of these patients were heterozygous ^51^. In our cohort, we had 250 patients with EOPD, and 5.6% (N = 14) of these patients carried a CNV on *PARK2*.

The role of homozygous and compound heterozygous variants, including CNVs on *PARK2* is well known, especially in EOPD ^5,15,45^. However, there is also increasing evidence that *PARK2* heterozygosity is a risk factor for PD and is associated with a decreased AAO ^45,46^. In our cohort, there was a significant association between the AAO and *PARK2* carrier status. Still, the role of heterozygous *PARK2* CNVs in altering PD susceptibility remains controversial ^16,52^. In order to correctly characterize PD patients, an integrated SNV-CNV analysis is needed, given the importance of both allele types for comprehensive genetic diagnosis in PD ^53^. Pankratz et al. showed that the frequency of carrying a single *PARK2* CNV was higher in PD patients compared to controls, while it was similar for carrying a single point mutation ^54^ Heterozygous *PARK2* CNV carrier status may still play a role in the development of PD despite its recessive inheritance ^46,55^, through a haploinsufficiency effect ^56^.

Some limitations of our study are that we did not validate all CNVs with a different method or sequence all *PARK2* CNV carriers. From our overall cohort (N = 1,379), a small portion were previously sequenced and/or CNV screened for other ongoing projects (see Supp. methods). Out of the 250 EOPD patients included in our analysis, 77 (30.8%) of them had been previously screened for CNVs with multiplex ligation-dependent probe amplification. This included seven of the 22 patients that were found in this current study carrying a CNV on *PARK2* or *SNCA*. Results for all 77 samples matched, confirming negatives and all seven positive carriers, showing that our CNV analysis pipeline was accurate and detected 100% of the known CNV carriers (with no false positives) found with a different method. In addition, out of the 14 EOPD patients who carried a *PARK2* CNV in our cohort, eight of them were also previously sequenced for *PARK2*. Based on data from the previous studies ^48,51^, out of the 14 EOPD patients who carried a *PARK2* CNV in our cohort, one would expect five to seven of them to be compound heterozygous for a *PARK2* mutation of any kind. We found three of them to be compound heterozygous, carrying both a CNV and a pathogenic SNV in *PARK2*, and one of these patients had a novel acceptor splice site mutation, reported previously ^57^. We also identified a fourth compound heterozygous sample through imputation (Supp. methods). Details for these four compound carriers are included in Supp. Table 2. This is very similar to the frequency reported in previous studies mentioned above^48,51^.

The large genetic variation in Latinos due to admixture from several populations (mostly European, Amerindian and African) creates a challenge when analyzing this population. For this reason, we established a workflow with rigorous quality control. We also constructed a Latino reference file from scratch for CNV calls, as publicly available reference files were all based on Europeans. In this study, we analyzed all samples together in order to boost statistical power. However, separate calling of CNVs in subpopulations based on admixture analysis is likely to yield more refined results. Thus, larger sample sizes will be needed to make discoveries specific to subpopulations of Latinos. Admixture mapping to examine the chromosomal location of the CNVs could also provide more insights about the relationship between PD genetics and ethnicity^58,59^.

To our knowledge, this is the first study that focuses on genome-wide CNVs in PD patients from Latin America. We believe that expanding the diversity of genetic studies for PD is necessary to understand the genetic profiles of these individuals and that our work will enrich current scientific knowledge about CNVs in this underrepresented population.

## Data Availability

All relevant data and methods are reported in the article and in the Supplement.

https://github.com/elifirem/LargePD_CNV

## Glossary

AAO = age at onset; BAF = B-allele frequency; CI = confidence interval; CNV = copy number variant; EOPD = early-onset Parkinson’s disease; HR = hazards ratio; Kb = kilobases; LRR = log R ratio; Mb = megabases; OR = odds ratio; PCA = principal component analysis; PD = Parkinson’s disease; QC = quality control; SNP = single nucleotide polymorphism; SNV = single nucleotide variant.

## Acknowledgments

We would like to thank all of the individuals that donated their samples as well as their time to participate in LARGE-PD, which made this and future projects possible. We would also like to thank all of our collaborators at the different Latin American sites for their efforts and support for building this incredible resource.

## Authors’ Roles

EIS and IFM designed the study. EIS, EPP, and LMN analyzed the data. IFM and DL supervised the study. EIS and IFM wrote the manuscript. All authors interpreted the data and revised the manuscript.

## Appendix

Members of the Latin American Research Consortium on the Genetics of PD (LARGE-PD): Argentina: Federico Micheli, Emilia Gatto.

Brazil: Vitor Tumas, Vanderci Borges, Henrique B. Ferraz, Carlos R.M. Rieder, Artur Shumacher-Schuh, Bruno L. Santos-Lobato.

Chile:Pedro Chaná.

Colombia: Carlos Velez-Pardo, Marlene Jimenez-Del-Rio, Francisco Lopera, Gonzalo Arboleda, Humberto Arboleda, Jorge Luis Orozco, Sonia Moreno, William Fernandez, Carlos E. Arboleda-Bustos.

Costa Rica:Jaime Fornaguera, Alvaro Hernández Guillén, Gabriel Torrealba Acosta.

Ecuador: Jorge Chang-Castello, Brennie Andreé Muñoz.

Honduras: Alex Medina, Anabelle Ferrera.

Mexico: Daniel Martinez-Ramirez, Mayela Rodriguez.

Peru: Mario Cornejo-Olivas, Pilar Mazzetti, Hugo Sarapura, Andrea Rivera, Luis Torres, Carlos Cosentino.

Puerto Rico: Angel Viñuela.

Uruguay: Elena Dieguez, Victor Raggio, Andres Lescano.

## Supplementary

**Supp. Table 1:** CNV characteristics. Profiles of CNVs called following QC steps in PD patients and controls.

|  | PD patients | Controls |
| --- | --- | --- |
| Number of samples | 747 | 632 |
| CNV carriers (%) | 692 (92.6%) | 582 (92.1%) |
| CNVs per sample | 6.9 | 5.2 |
| Number of CNVs | 5,155 | 3,257 |
| Duplications | 2,984 | 1,538 |
| Deletions | 2,171 | 1,719 |
| Mean size of CNVs (Kb) | 99 | 102 |
| Median size of CNVs (Kb) | 52 | 55 |
| Mean number of SNPs per CNV | 111 | 94 |
| Median Quality Score of duplications | 0.967 | 0.966 |
| Median Quality Score of deletions | -0.995 | -0.994 |
| Out CNVs | 133,462 | 100,678 |
| Duplications | 45,548 | 20,268 |
| Deletions | 87,914 | 80,410 |

PD: Parkinson's disease, CNV: copy number variant, Kb: kilobases.

**Supp. Table 2:** Compound heterozygous PD patients.

|  | **Age at Visit** | **Age at Onset** | **Country** | ***PARK2* CNV** | ***PARK2* SNV** |
| --- | --- | --- | --- | --- | --- |
| Patient 1 | 40 | 25 | Colombia | Duplication (exon 3) | rs137853058 - p.Cys212Tyr |
| Patient 2<sup>a</sup> | 47 | 15 | Peru | Deletion (exon 7) | IVS5-1G>A - splice site |
| Patient 3 | 24 | 12 | Uruguay | Deletion (exons 3-6) | rs746646126 - p.Trp74CysfsTer8<sup>b</sup> |
| Patient 4 | 59 | 41 | Brazil | Deletion (exon 3) | rs754809877 - p.Asn52MetfsTer29<sup>c</sup> |

CNV: copy number variant, SNV: single nucleotide variant.
<sup>a</sup> This patient and his family have been previously described <sup>57</sup>
<sup>b</sup> This patient is homozygous for this variant
<sup>c</sup> Imputed using TopMed

**Supp. Figure 1:**
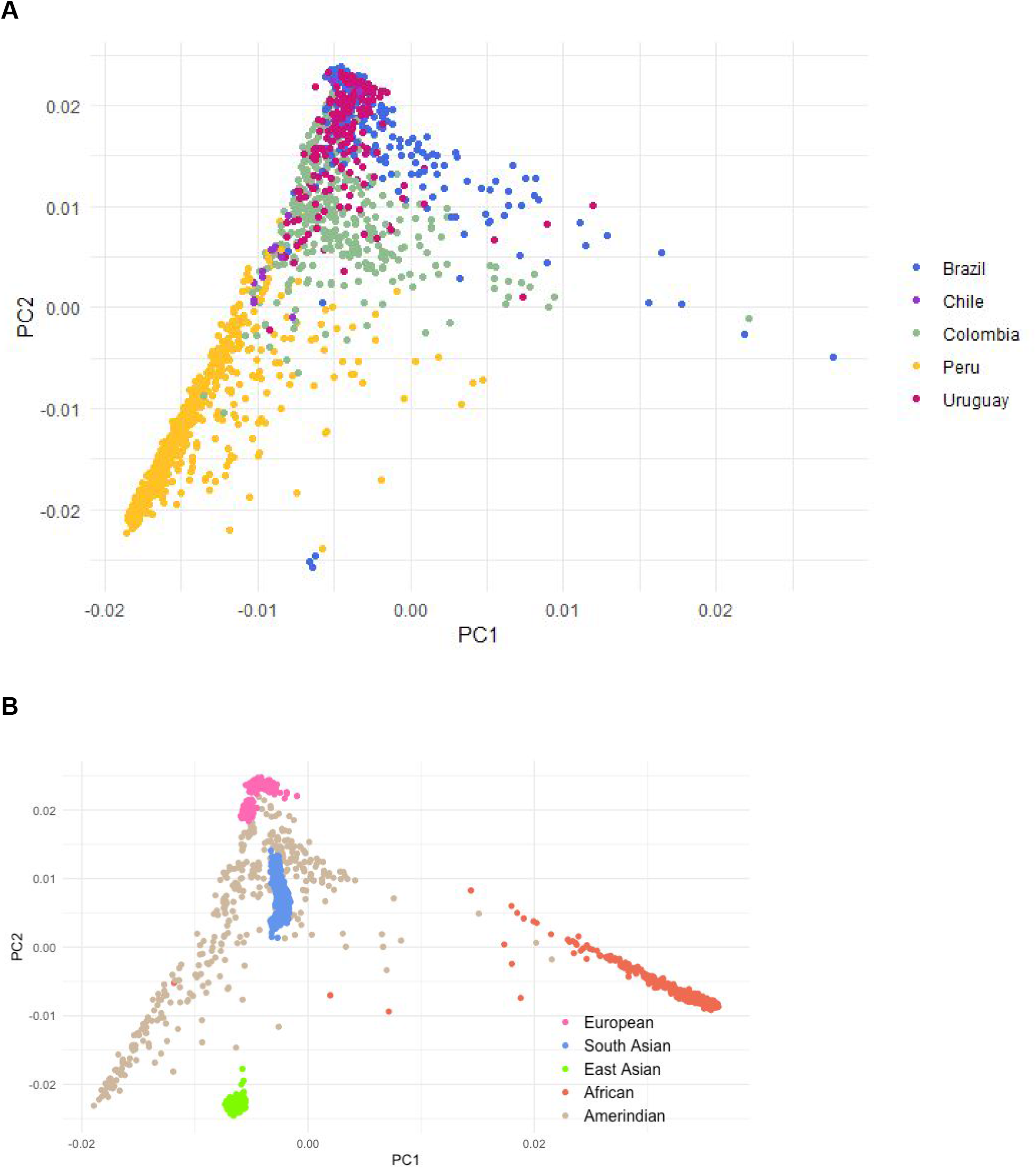
(A) PCA of LARGE-PD individuals included in the study, color-coded by country of origin. (B) PCA of 1000 Genomes population for comparison, color-coded by different populations.

**Supp. Figure 2:**
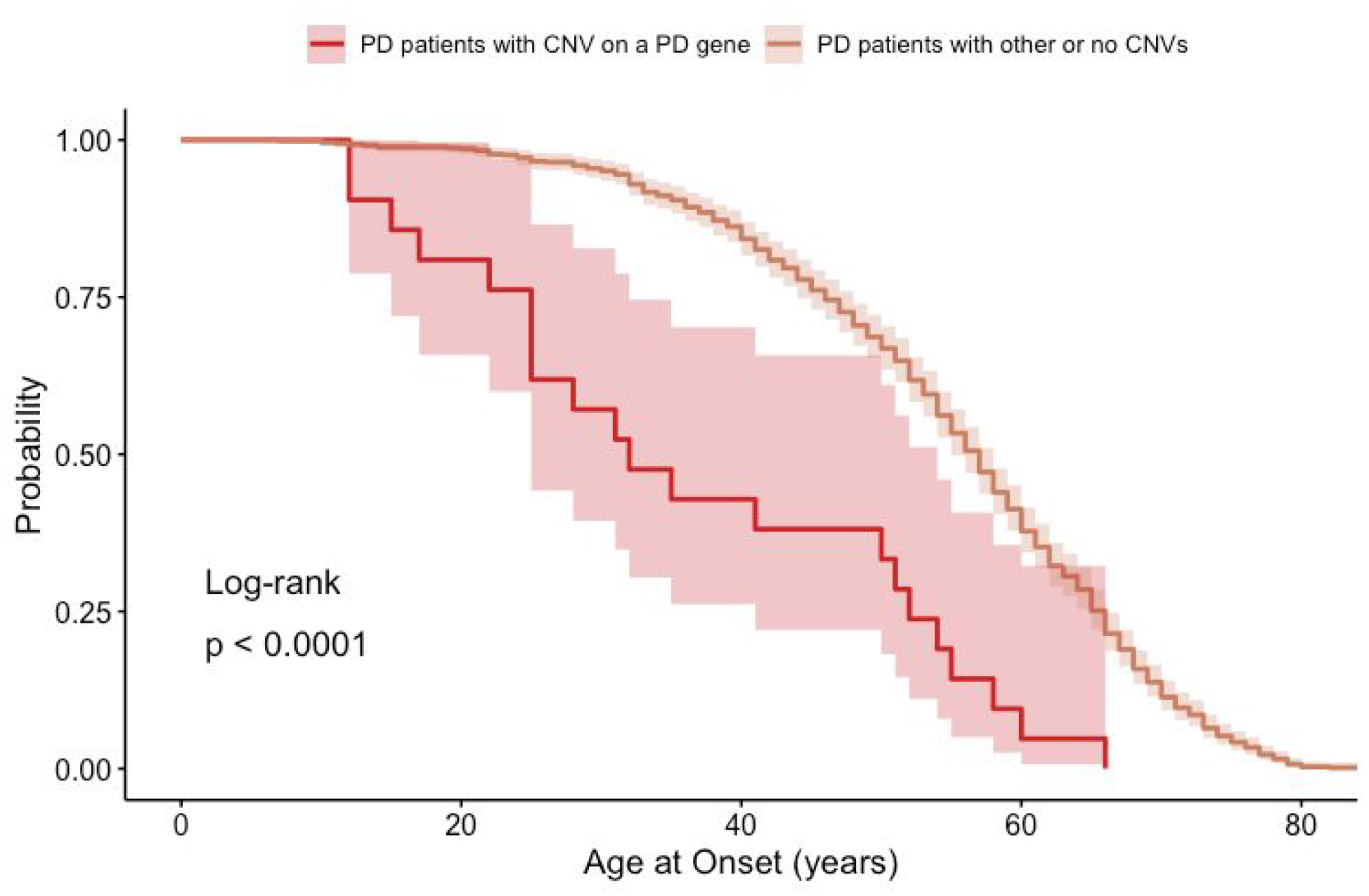
Kaplan-Meier estimates of PD patients carrying a CNV on a known PD gene and patients with other or no CNVs. Probability: probability of not having symptoms associated with PD. Age at Onset: time to onset of PD symptoms. Highlight around the curves shows 95% confidence intervals.

**Supp. Methods:**

From our overall cohort (N = 1,379), 117 (8.4%) of the individuals were previously sequenced and/or CNV screened for other ongoing projects. Forty of the samples with self-reported family history were capture-sequenced for all coding regions using a custom neurodegenerative panel which included 21 PD related genes: *ATP13A2, ATP1A3, DNAJC6, DNAJC13, EIF4G1, FBXO7, GCH1, GBA, GIGYF2, HTRA2, LRRK2, PARK2, PARK7, PINK1, PLA2G6, RAB39B, SLC6A3, SNCA, SNCB, SYNJ1, TAF1, TARDBP, TMEM230, VPS13C*, and *VPS35*. Seventy-seven patients with EOPD were Sanger sequenced and CNV screened using multiplex ligation-dependent probe amplification for *PARK2 and SNCA*. We also imputed LARGE-PD dataset (807 PD patients and 690 controls) using the Trans-Omics for Precision Medicine (TOPMed) Project ^60^ which identified a fourth compound heterozygous individual.

## References

1. de Lau, L. M. & Breteler, M. M. Epidemiology of Parkinson’s disease. Lancet Neurol. 5, 525–535 (2006).

2. Global, regional, and national burden of Parkinson’s disease, 1990–2016: a systematic analysis for the Global Burden of Disease Study 2016. Lancet Neurol. 17, 939–953 (2018).

3. Noyce, A. J. et al. Meta-analysis of early nonmotor features and risk factors for Parkinson disease. Ann. Neurol. 72, 893–901 (2012).

4. Singleton, A. B., Farrer, M. J. & Bonifati, V. The genetics of Parkinson’s disease: progress and therapeutic implications. Mov. Disord. Off. J. Mov. Disord. Soc. 28, 14–23 (2013).

5. Domingo, A. & Klein, C. Chapter 14 - Genetics of Parkinson disease. in Handbook of Clinical Neurology (eds. Geschwind, D. H., Paulson, H. L. & Klein, C.) vol. 147 211–227 (Elsevier, 2018).

6. Klein, C. & Westenberger, A. Genetics of Parkinson’s Disease. Cold Spring Harb. Perspect. Med. 2, (2012).

7. Polymeropoulos, M. H. et al. Mutation in the α-Synuclein Gene Identified in Families with Parkinson’s Disease. Science 276, 2045–2047 (1997).

8. Krüger, R. et al. Ala30Pro mutation in the gene encoding alpha-synuclein in Parkinson’s disease. Nat. Genet. 18, 106–108 (1998).

9. Zarranz, J. J. et al. The new mutation, E46K, of alpha-synuclein causes Parkinson and Lewy body dementia. Ann. Neurol. 55, 164–173 (2004).

10. Appel-Cresswell, S. et al. Alpha-synuclein p.H50Q, a novel pathogenic mutation for Parkinson’s disease. Mov. Disord. Off. J. Mov. Disord. Soc. 28, 811–813 (2013).

11. Lesage, S. et al. G51D α-synuclein mutation causes a novel parkinsonian-pyramidal syndrome. Ann. Neurol. 73, 459–471 (2013).

12. Singleton, A. B. et al. alpha-Synuclein locus triplication causes Parkinson’s disease. Science 302, 841 (2003).

13. Ibáñez, P. et al. Causal relation between alpha-synuclein gene duplication and familial Parkinson’s disease. Lancet Lond. Engl. 364, 1169–1171 (2004).

14. Chartier-Harlin, M.-C. et al. Alpha-synuclein locus duplication as a cause of familial Parkinson’s disease. Lancet Lond. Engl. 364, 1167–1169 (2004).

15. Kitada, T. et al. Mutations in the parkin gene cause autosomal recessive juvenile parkinsonism. Nature 392, 605–608 (1998).

16. Mata, I. F., Lockhart, P. J. & Farrer, M. J. Parkin genetics: one model for Parkinson’s disease. Hum. Mol. Genet. 13, R127–R133 (2004).

17. Pankratz, N. et al. Copy Number Variation in Familial Parkinson Disease. PLoS ONE 6, (2011).

18. Kim, J.-S. et al. Comparative genome hybridization array analysis for sporadic Parkinson’s disease. Int. J. Neurosci. 118, 1331–1345 (2008).

19. Liu, X. et al. Increased Rate of Sporadic and Recurrent Rare Genic Copy Number Variants in Parkinson’s Disease Among Ashkenazi Jews. Mol. Genet. Genomic Med. 1, 142–154 (2013).

20. Darvish, H. et al. Detection of copy number changes in genes associated with Parkinson’s disease in Iranian patients. Neurosci. Lett. 551, 75–78 (2013).

21. Toft, M. & Ross, O. A. Copy number variation in Parkinson’s disease. Genome Med. 2, 62 (2010).

22. Ibáñez, P. et al. α-Synuclein Gene Rearrangements in Dominantly Inherited Parkinsonism: Frequency, Phenotype, and Mechanisms. Arch. Neurol. 66, 102–108 (2009).

23. Bureau, U. C. Projections of the Size and Composition of the U.S: 2014–2060. The United States Census Bureau https://www.census.gov/library/publications/2015/demo/p25-1143.html.

24. Popejoy, A. B. & Fullerton, S. M. Genomics is failing on diversity. Nature 538, 161 (2016).

25. Van Den Eeden, S. K. et al. Incidence of Parkinson’s Disease: Variation by Age, Gender, and Race/Ethnicity. Am. J. Epidemiol. 157, 1015–1022 (2003).

26. Wright Willis, A., Evanoff, B. A., Lian, M., Criswell, S. R. & Racette, B. A. Geographic and Ethnic Variation in Parkinson Disease: A Population-Based Study of US Medicare Beneficiaries. Neuroepidemiology 34, 143–151 (2010).

27. Zabetian, C. P. & Mata, I. F. LARGE-PD: Examining the genetics of Parkinson’s disease in Latin America. Mov. Disord. 32, 1330–1331 (2017).

28. Gibb, W. R. & Lees, A. J. The relevance of the Lewy body to the pathogenesis of idiopathic Parkinson’s disease. J. Neurol. Neurosurg. Psychiatry 51, 745–752 (1988).

29. Chang, C. C. et al. Second-generation PLINK: rising to the challenge of larger and richer datasets. GigaScience 4, (2015).

30. Niestroj, L.-M. et al. Evaluation of copy number burden in specific epilepsy types from a genome-wide study of 18,564 subjects. http://biorxiv.org/lookup/doi/10.1101/651299 (2019) doi:10.1101/651299.

31. A global reference for human genetic variation. Nature 526, 68–74 (2015).

32. Wang, K. et al. PennCNV: An integrated hidden Markov model designed for high-resolution copy number variation detection in whole-genome SNP genotyping data. Genome Res. 17, 1665–1674 (2007).

33. Manichaikul, A. et al. Robust relationship inference in genome-wide association studies. Bioinforma. Oxf. Engl. 26, 2867–2873 (2010).

34. Huang, A. Y. et al. Rare Copy Number Variants in NRXN1 and CNTN6 Increase Risk for Tourette Syndrome. Neuron 94, 1101–1111. e7 (2017).

35. Macé, A. et al. New quality measure for SNP array based CNV detection. Bioinformatics 32, 3298–3305 (2016).

36. Ensembl 2019 | Nucleic Acids Research | Oxford Academic. https://academic.oup.com/nar/article/47/D1/D745/5165265.

37. Quinlan, A. R. & Hall, I. M. BEDTools: a flexible suite of utilities for comparing genomic features. Bioinforma. Oxf. Engl. 26, 841–842 (2010).

38. Bandres-Ciga, S., Diez-Fairen, M., Kim, J. J. & Singleton, A. B. Genetics of Parkinson’s disease: An introspection of its journey towards precision medicine. Neurobiol. Dis. 137, 104782 (2020).

39. Lunati, A., Lesage, S. & Brice, A. The genetic landscape of Parkinson’s disease. Rev. Neurol. (Paris) 174, 628–643 (2018).

40. SurajGupta/r-source. GitHub https://github.com/SurajGupta/r-source.

41. R: The R Project for Statistical Computing. https://www.r-project.org/.

42. Therneau, T. M., until 2009), T. L. (original S.->R port and R. maintainer, Elizabeth, A. & Cynthia, C. survival: Survival Analysis. (2020).

43. Redon, R. et al. Global variation in copy number in the human genome. Nature 444, 444 (2006).

44. Jakobsson, M. et al. Genotype, haplotype and copy-number variation in worldwide human populations. Nature 451, 998–1003 (2008).

45. Lücking, C. B. et al. Association between early-onset Parkinson’s disease and mutations in the parkin gene. N. Engl. J. Med. 342, 1560–1567 (2000).

46. Sun, M. et al. Influence of Heterozygosity for Parkin Mutation on Onset Age in Familial Parkinson Disease: The GenePD Study. Arch. Neurol. S3, 826–832 (2006).

47. Oliveira, S. A. et al. Parkin mutations and susceptibility alleles in late-onset Parkinson’s disease. Ann. Neurol. 53, 624–629 (2003).

48. Marder, K. S. et al. Predictors of parkin mutations in early-onset Parkinson disease: the consortium on risk for early-onset Parkinson disease study. Arch. Neurol. B 7, 731–738 (2010).

49. Hedrich, K. et al. Evaluation of 50 probands with early-onset Parkinson’s disease for Parkin mutations. Neurology 58, 1239–1246 (2002).

50. Periquet, M. et al. Parkin mutations are frequent in patients with isolated early-onset parkinsonism. Brain 12B, 1271–1278 (2003).

51. Camacho, J. L. G. et al. High frequency of Parkin exon rearrangements in Mexican-mestizo patients with early-onset Parkinson’s disease. Mov. Disord. 27, 1047–1051 (2012).

52. Yu, E. et al. Analysis of heterozygous PRKN variants and copy number variations in Parkinson’s disease. *medRxiv* 2020.05.07.20072728 (2020) doi: 10.1101/2020.05.07.20072728.

53. Robak, L. A. et al. Integrated Sequencing & Array Comparative Genomic Hybridization in Familial Parkinson’s Disease. http://biorxiv.org/lookup/doi/10.1101/828566 (2019) doi:10.1101/828566.

54. Pankratz, N. et al. Parkin dosage mutations have greater pathogenicity in familial PD than simple sequence mutations. Neurology 73, 279–286 (2009).

55. Huttenlocher, J. et al. Heterozygote carriers for CNVs in PARK2 are at increased risk of Parkinson’s disease. Hum. Mol. Genet. 24, 5637–5643 (2015).

56. Klein, C., Lohmann-Hedrich, K., Rogaeva, E., Schlossmacher, M. G. & Lang, A. E. Deciphering the role of heterozygous mutations in genes associated with parkinsonism. Lancet Neurol. 6, 652–662 (2007).

57. Cornejo-Olivas, M. et al. A Peruvian family with a novel PARK2 mutation: Clinical and Pathological Characteristics. Parkinsonism Relat. Disord. 21, 444–448 (2015).

58. Genovese, G. et al. Using population admixture to help complete maps of the human genome. Nat. Genet. 45, 406–414e2 (2013).

59. Lou, H. et al. Copy number variations and genetic admixtures in three Xinjiang ethnic minority groups. Eur. J. Hum. Genet. 23, 536–542 (2015).

60. Taliun, D. et al. Sequencing of 53,831 diverse genomes from the NHLBI TOPMed Program. *bioRxiv* 563866 (2019) doi:10.1101/563866.

